# Social and Mental Health Pathways to Institutional Trust: A Cohort Study

**DOI:** 10.1101/2024.12.05.24318551

**Authors:** Vincent Paquin, Diana Miconi, Samantha Aversa, Janique Johnson-Lafleur, Sylvana Côté, Marie-Claude Geoffroy, Sinan Gülöksüz

## Abstract

**Objective:** Trust in institutions such as the government is lower in the context of mental health problems and socio-economic disadvantage. However, the roles of structural inequality, interpersonal factors, and mental health on institutional trust remain unclear. This study aimed to examine the associations of social and mental health factors, from early life to adulthood, with institutional trust.

**Method:** Participants (n=1347; 57.2% female) were from the population-based Québec Longitudinal Study of Child Development (1997-2021). Trust in 13 institutions was self-reported at age 23. Predictors were 20 social and mental health factors during early life, adolescence, and adulthood. Associations were examined with linear regressions corrected for false discovery rate. Pathways were explored using the temporal Peter-Clark algorithm.

**Results:** Early-life factors associated with lower levels of trust were male sex, racialized minority status, low household income, and maternal history of depression and antisocial behaviors. After adjusting for early-life factors, adolescence factors associated with lower levels of trust were internalizing and externalizing problems, bullying exposure, and school difficulties. Independently of early-life or adolescence factors, adulthood factors associated with lower levels of trust were perceived stress, psychotic experiences, suicidal ideas, and seeking professional help, whereas greater social capital was associated with greater trust. Temporal Peter-Clark analyses identified social capital and psychotic experiences as potential proximal determinants of institutional trust.

**Conclusion:** This study identified factors associated with institutional trust reflecting an interplay between structural inequality, interpersonal relationships, and mental health over development. Interventions aimed at promoting social inclusion may improve institutional trust and population wellbeing.

## INTRODUCTION

Trust in institutions is the belief that institutions, such as the government, health care, or media, will act in a favorable way towards oneself (Bauer & Freitag, 2018). Institutional trust is included in many government-led surveys worldwide as an indicator of population wellbeing and social cohesion (Uslaner, 2018). Yet during recent years, trust in institutions has been low in many countries such as Canada and the U.S, and its possible decline has been attributed to economic uncertainties, the COVID-19 pandemic, and polarizing effects of social media (H. E. Brady & Kent, 2022; Lorenz-Spreen et al., 2023; Steinburg, 2024). Trust in institutions is a key issue for health care and particularly mental health care: not only can institutional mistrust hamper the accessibility and reach of mental health institutions (Gulliver et al., 2010), but population surveys also indicate an association between lower levels of trust and poorer self-reported mental health (Buck-McFadyen et al., 2019; Economou et al., 2014; Hudson, 2006; Lindstrom & Mohseni, 2009), suggesting that mistrust or its underlying causes could be a risk factor for mental health problems. Therefore, understanding the pathways to institutional (mis)trust over development could guide population-based approaches aimed at promoting mental health. Drawing from psychological and sociological theories of trust, we propose a developmental and prospective investigation of institutional trust that focuses on three broad categories of factors: structural inequality, interpersonal relationships, and mental health.

Psychological and sociological theories offer complementary explanations to the development of institutional trust over the life course. The psychological propensity model (Glanville & Paxton, 2007) and the familial socialization of trust hypothesis (Uslaner, 2002) posit that individuals acquire a general propensity to trust or distrust early in life through interactions with their caregivers. The notion of a general propensity for trust is supported by positive associations between social and political trust (Newton & Zmerli, 2011), and its familial socialization is supported by evidence of a longitudinal association between early-life adversity and adolescent social trust (Reiter et al., 2023). Putnam’s theory of social capital advances that individuals acquire norms of trust and reciprocity through their relationships with other members of society, and these norms shape their willingness to engage with institutions (Putnam, 1993). In support of this model, some studies (de Vroome et al., 2013; Kääriäinen, 2007), but not all (Meeusen et al., 2024), have found cross-sectional associations between indicators of social capital (e.g., frequency of contact with other people) and trust in the government and police. Performance theory contends that the level of institutional of trust depends on individual experiences of the institution’s capacity to produce favorable outcomes (Van Craen, 2013). For example, unfair treatment of racialized minority groups by the police could explain their lower levels of trust in this institution (Murphy et al., 2022; Wilkes & Wu, 2018). Informational habitus, i.e., the habitual ways of individuals of accessing and using information (Vivion & Malo, 2023) could further influence how individuals appraise the performance of institutions. A recent review highlights cross-sectional evidence for a link between internet use and lower levels of trust in political institutions and news media, possibly reflecting the effect of polarizing online contents and, in the context of authoritarian regimes, freer circulation of anti-government information (Lorenz-Spreen et al., 2023).

These theories of trust point to a range of social and psychological factors shaping institutional trust over development. However, as illustrated above, the corresponding empirical evidence is mostly cross-sectional and tends to focus on small sets of risk factors. Given the potential intercorrelations among factors such as socio-economic disadvantage, early-life adversity, social capital, informational habitus, and other determinants of trust, the relative contribution of each factor over the life course remains unclear. A broader examination of the development of institutional (mis)trust, across multiple potential risk factors and in a prospective fashion, could advance theories and interventions on institutional trust by highlighting key pathways involved in the formation of trust.

Social approaches to mental health emphasize the interplay of structural inequality, social relationships, and mental health problems (Kirmayer, 2024), three factors that in theory would be expected to influence institutional trust. Structural inequality refers to disparities in resources that result from discriminatory treatment from institutions, and which affect specific groups in a given society (Assari, 2019). Following performance theory, discriminatory treatment makes institutions less trustworthy for affected groups. For instance, groups in the United States who have reported lower levels of trust in institutions include lower-income (Newton et al., 2018), Black (Macdonald & Stokes, 2006), and gender diverse individuals (Mason et al., 2024). A study in the United Kingdom found a prospective association between socio-economic disadvantage at birth and lower levels of political trust in adulthood (Schoon & Cheng, 2011). Interpersonal relationships, including the early-life family environment and later social capital, may also influence institutional trust: experiences of reciprocity during development are thought to shape how individuals more broadly infer the trustworthiness of society, including its institutions (Newton et al., 2018; Sturgis et al., 2015). Greater use of social media may serve to nurture social relationships but, as a part of a person’s informational habitus, may also contribute to lower levels of institutional trust through access to polarizing or unfavorable news (W. J. Brady et al., 2023; Lorenz-Spreen et al., 2023). Lastly, mental health problems can be interrelated with institutional trust at the social and psychological levels: mental health problems expose individuals to stigma, which contributes to structural inequality and social exclusion (Thornicroft et al., 2022), and mental health problems such as psychotic experiences may emerge from or reflect an underlying propensity for general mistrust (Lopes et al., 2020).

Using a large population-based prospective cohort, this study therefore aimed to examine the social and mental health pathways to institutional trust from early life to adulthood. We made the general hypothesis that factors related to structural inequality, interpersonal problems, and mental health problems would be associated with lower levels of trust.

## METHODS

### Participants

We analyzed data from the Québec Longitudinal Study of Child Development, a population-based cohort of 2120 individuals born in the Province of Québec, Canada between 1997 and 1998 (https://www.jesuisjeserai.stat.gouv.qc.ca/) (Orri et al., 2020). Participants and their parents were recruited through stratified sampling from birth registries. Data collection covered all regions, except for Cree and Inuit territories and Indigenous communities due to differences in institutional services and socio-cultural context. Participants were followed annually or biennially from five months to 23 years of age (1997-2021). The cohort was conducted by the Institut de la statistique du Québec and approved by the ethics committees of the Institut de la statistique du Québec, Sainte-Justine Hospital Research Centre, and Montreal West Island Integrated University Health and Social Services Centre. Informed written consent, assent, or both were obtained for each data collection. This study followed the Strengthening the Reporting of Observational Studies in Epidemiology (STROBE) reporting guideline (Vandenbroucke et al., 2007). Analyses were conducted between January and October 2024.

### Institutional trust

Institutional trust was measured at age 23 years in 2021 with a 13-item self-report questionnaire adapted from the Canadian and U.S. General Social Surveys (Schellenberg, 2004). Participants were asked “How much confidence do you have in…” for various institutions (e.g., the government, healthcare, media; see Figure 1) on a scale of 1=“A great deal of confidence” to 4=“No confidence at all”. Answers were reverse coded so that a higher score indicates greater trust. To our knowledge, the structural validity of this questionnaire has not been examined before, but previous research has found cross-sectional associations of similar items with greater psychological distress and poorer self-rated mental health (Achdut, 2023; Buck-McFadyen et al., 2019; Lindstrom & Mohseni, 2009). Here, internal reliability was excellent (α=0.92; ω_total_=0.94), and exploratory and confirmatory factor analyses identified three factors within the scale: trust in social, economic, and mediatic institutions (Figure 1 and Note S1 in the Supplementary Material).

**Figure 1.**
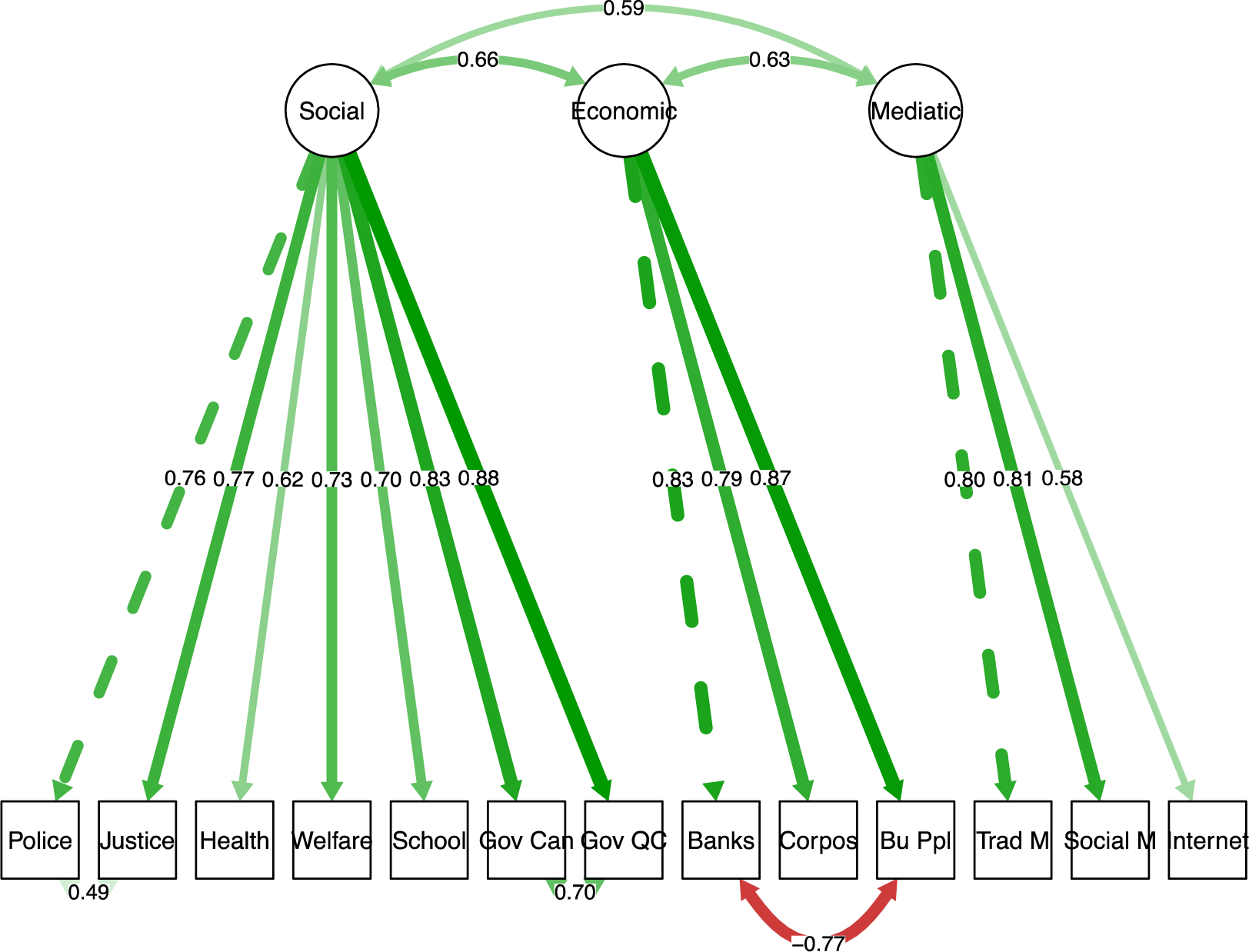
Identification of three domains of institutional trust through factor analysis. Confirmatory factor analysis in half of the sample (n=674). From left to right, the items are the police, justice system and courts, health care system, welfare system, school system, federal parliament (of Canada), provincial parliament (of Québec), banks, major corporations, businesspeople, traditional media, social media, and internet. Data compiled from the Québec Longitudinal Study of Child Development (1998– 2021), Gouvernement du Québec, Institut de la statistique du Québec.

### Social and mental health factors

Measures were collected across three developmental periods: early-life (birth to 29 months of age), adolescence (15 years), and adulthood (23 years, concurrently with the measure of institutional trust). Time points and variables were selected based on data availability, theory, and parsimony. We selected variables related to structural inequality, interpersonal factors, mental health problems, or their antecedents, such as socio-demographic factors and parental mental health. For mental health problems, we prioritized measures that cut across diagnostic categories and levels of severity. Note S2 in the Supplementary Material provides more information on these decisions.

Table S1 in the Supplementary Material details the source and content of each measure. Briefly, early-life factors were sex assigned at birth, racialized minority status (defined as parental identification of the child as Arab or West Asian, Asian, Black, Indigenous, Latinx, Mixed, or other ethnoracial groups other than White), low household income (relative to regional thresholds; Desrosiers et al., 2001), parental history of depression, and parental history of antisocial behaviors. Adolescence factors included maternal reports of neighborhood trust (i.e., trustworthiness of neighborhood residents) and family cohesion, and participant reports of school difficulties, exposure to bullying, and internalizing and externalizing problems. Adulthood factors were self-reported and included gender diverse status (i.e., transgender or other non-cisgender identification), perceived stress, psychotic experiences, suicidal ideas in the past year, seeking professional help for mental health in the past year, social capital (i.e., connection to others and availability of social support), and social media use (i.e., daily time spent on social media).

### Statistical analysis

Analyses were conducted in R version 4.3.1 (R Foundation for Statistical Computing) in participants with complete data on institutional trust. Of the cohort, 1368 participants completed the assessment at age 23 years, and 1347 (771 [57.2%] female) had data on institutional trust. Within the analytic sample of n=1347, data availability ranged between 87.1%–100%, except for paternal history of depression, neighborhood trust, and family cohesion which had more missing data (71.9%–83.8%; Table S2 in the Supplementary Material). Characteristics of participants included in and excluded from analyses on the basis of missing data were compared using effect sizes. Included participants were more likely to be female and in households with sufficient income and had mothers who were more educated (effect sizes > 0.100; Table S3 in the Supplementary Material).

To examine associations of social and mental health factors with institutional trust, we conducted linear regression models for each predictor. Sex was included as a covariate in all models, and all predictors from the preceding timepoint were additionally included as covariates: early-life variables were covariates for adolescence predictors, and adolescence variables were covariates for adulthood predictors. To account for the testing of multiple predictors within each time period, statistical significance was defined as p<.05 after correction for false discovery rate using the Benjamini & Hochberg (1995) method. Missing data on predictors was replaced with multiple imputation using chained equations (Buuren & Groothuis-Oudshoorn, 2011): 40 datasets were generated based on a large set of predictor variables, which were selected procedurally according to a criterion of *r*>0.15 with the imputed variable (Buuren & Groothuis-Oudshoorn, 2011; van Buuren et al., 1999).

To identify pathways of association among all variables including institutional trust, we used the temporal Peter-Clark (tPC) algorithm (Petersen et al., 2021; Spirtes & Glymour, 1991). The tPC algorithm is a data-driven approach to uncover potential causal pathways in the form of a directed acyclic graph. The algorithm starts with a fully connected graph. Edges between variables are removed if their association can be explained by any set of covariables that are temporally concurrent or anterior. The impact of covariables is determined by conditional independence testing and using a pre-defined sparsity level, meaning that with a lower sparsity level, only relationships that more robustly survive adjustment for other variables are retained in the graph. Then, edges between variables are oriented, when possible, based on logical rules (e.g., the identification of v-structures and colliders; Petersen et al., 2021; Spirtes & Glymour, 1991) and the temporality of variables. This algorithm has been shown to be mathematically correct for recovering causal pathways, but it relies on strong assumptions, notably that there are no unobserved confounders (Spirtes & Glymour, 1991). We applied the tPC algorithm with 4 sparsity levels: 0.01, 0.001, 0.0001, and 0.00001 (Petersen et al., 2021). Missing data was handled with test-wise deletion since it is less computationally expensive and performs similarly to multiple imputation when there is a mix of Gaussian and discrete variables (Witte et al., 2022). We performed 100 bootstraps per model to estimate the stability of edges and interpreted stability as modest if edges were identified in 50-74% of bootstraps and good if they were identified in 75-100% (Foraita et al., 2024; Petersen et al., 2023).

We interpreted results with a primary interest in potential causal effects. Following calls for greater clarity on causal inference in health research (Hernán, 2018), we used terms such as “putative effects” to transparently communicate our interpretation of causality, while considering limitations to causal inference and alternative possibilities in the Discussion.

## RESULTS

### Associations with institutional trust

Descriptive statistics on key study variables are presented in Table 1. Early-life factors significantly associated with lower levels of institutional trust were racialized minority status, low household income, maternal history of depression, and maternal history of antisocial behaviors. Female sex was associated with greater institutional trust. Adolescence factors associated with lower levels of institutional trust were exposure to bullying, school difficulties, and internalizing and externalizing problems. Adulthood factors associated with lower levels of institutional trust were higher levels of perceived stress and psychotic-like experiences, suicidal ideas, and seeking professional help. Greater social capital was associated with greater institutional trust. Associations were generally consistent across the three domains of institutional trust (social, political, and mediatic factors) and alternative sets of covariables (Figures S1-S6).

**Table 1.** Descriptive statistics of key variables in the study.

| Variable | N (%) or Mean (SD) |
| --- | --- |
| *Early life (0-2.5 years)* |  |
| Sex assigned at birth |  |
| Male | 576 (42.8%) |
| Female | 771 (57.2%) |
| Ethnoracial groups |  |
| Arab or West Asian | 15 (1.12%) |
| Asian | 15 (1.12%) |
| Black | 24 (1.79%) |
| Mixed | 24 (1.79%) |
| White | 1234 (91.8%) |
| Other (Indigenous, Latinx, or “Other”) | 32 (2.38%) |
| Low household income |  |
| Yes | 1063 (79.8%) |
| No | 269 (20.2%) |
| Maternal history of depression |  |
| Yes | 289 (21.7%) |
| No | 1040 (78.3%) |
| Paternal history of depression |  |
| Yes | 156 (14.1%) |
| No | 952 (85.9%) |
| Maternal adolescent antisocial behaviors, range: 0-5 | 0.81 (0.92) |
| Paternal adolescent antisocial behaviors, range: 0-5 | 0.64 (0.91) |
| *Adolescence (15 years)* |  |
| Neighborhood trust, range: 1-4 | 3.28 (0.56) |
| Family cohesion, range: 7-28 | 24.1 (3.04) |
| Bullying exposure, range: 6-18 | 8.20 (2.59) |
| School difficulties, range: 0-1 | 0.24 (0.26) |
| Internalizing problems, range: 0-10 | 3.43 (1.91) |
| Externalizing problems, range: 0-10 | 1.43 (0.89) |
| ***Adulthood (23 years)*** |  |
| Gender diverse status |  |
| Yes | 30 (2.23%) |
| No | 1317 (97.8%) |
| Perceived stress, past month, range: 0-40 | 17.9 (7.20) |
| Psychotic experiences, lifetime occurrence, range: 15-60 | 18.9 (4.37) |
| Suicidal ideas, past year |  |
| Yes | 112 (8.48) |
| No | 1209 (91.5%) |
| Help seeking for mental health, past year |  |
| Yes | 276 (20.5%) |
| No | 1071 (79.5%) |
| Social capital, range: 0-40 | 27.0 (3.92) |
| Social media use, hours/day, range: 0-6 | 2.67 (1.74) |
| Institutional trust, range: 13-52 | 34.4 (6.89) |

**Figure 2.**
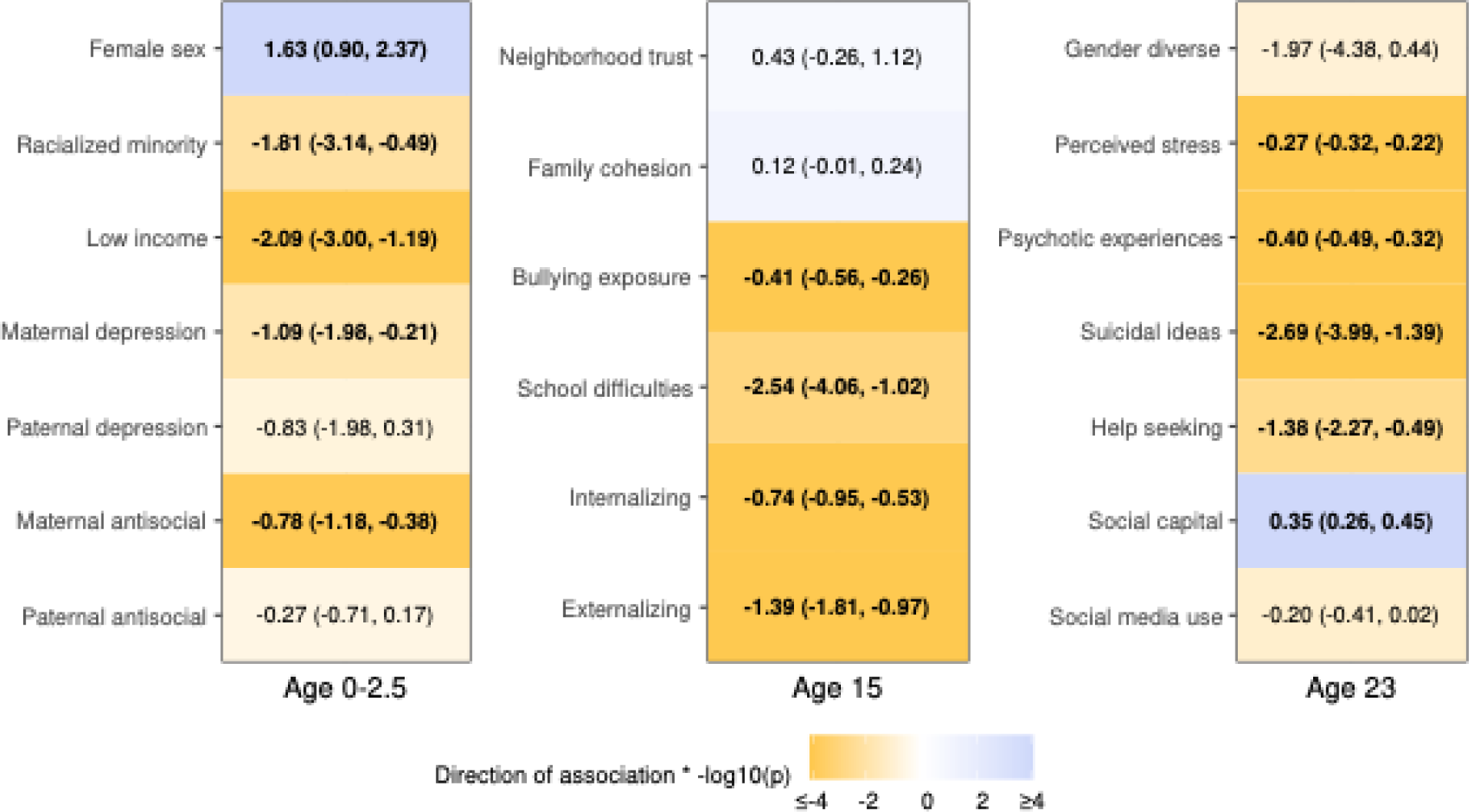
Associations of social and mental health factors with institutional trust. Unstandardized regression coefficients (95% confidence intervals) pooled over 40 multiply imputed datasets (n=1347). Control variables were sex for early-life factors (age 0-2.5 years), sex and early-life factors for adolescence factors (age 15 years), and sex and adolescence factors for adulthood factors (age 23 years). Bold indicates p<.05 after adjustment for false discovery rate across columns. Darker shades indicate smaller p-values (orange: negative association; blue: positive association). Data compiled from the Québec Longitudinal Study of Child Development (1998–2021), Gouvernement du Québec, Institut de la statistique du Québec.

### Pathways to institutional trust

We focused here on the most parsimonious model (sparsity level=0.00001) (Figure 3). In this model, there were putative effects of psychotic experiences and social capital on institutional trust. Institutional trust had a putative effect on perceived stress. Externalizing problems had an indirect effect on institutional trust via psychotic experiences. Bullying exposure shared an undirected edge with externalizing problems. Other factors were not linked to institutional trust.

Across bootstraps, edges involving institutional trust were stable, but their orientations were not (Table S3). The edge between psychotic experiences and institutional trust was found in 72% of models: orientations were variably from psychotic experiences to institutional trust (34%), from institutional trust to psychotic experiences (28%), or undirected (10%). The edge between social capital and institutional trust was found in 77% of models: orientations were from social capital to institutional trust (24%), from institutional trust to social capital (10%), and undirected (43%).

Models with more lenient sparsity levels (Figures S7-S9) also found putative effects of psychotic experiences on institutional trust. Putative effects of social capital on institutional trust were less consistent: they were found in one model (sparsity=0.01) and were the reverse (from institutional trust to social capital) in the other two models. Bootstrap stability was similar across sparsity levels (see Supplementary Data at https://osf.io/82f4q/?view_only=96a7f46060c84a0a8cd9e995c0655afe).

**Figure 3.**
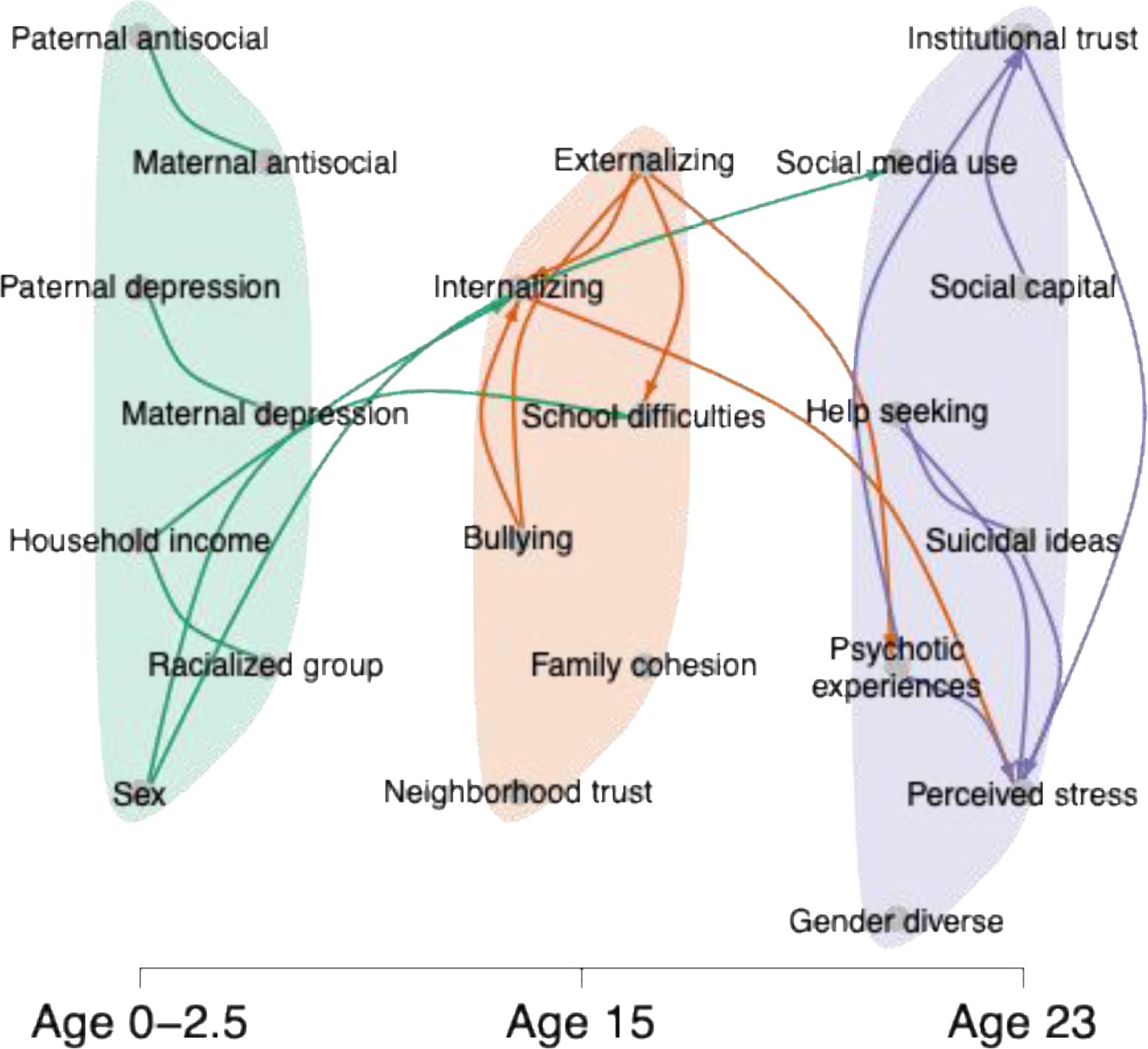
Paths between social factors, mental health, and institutional trust (sparsity=0.00001) Temporal Peter-Clark algorithm with test-wide deletion (n=1347). Edges (lines) indicate putative causal relationships between variables. Some edges have no arrowheads, indicating that the direction of causality could not be determined by the algorithm. Data compiled from the Québec Longitudinal Study of Child Development (1998–2021), Gouvernement du Québec, Institut de la statistique du Québec.

## DISCUSSION

### Social factors and institutional trust

Putative effects of structural inequality on institutional trust were supported by the associations of low household income, racialized minority status and school difficulties with lower levels of trust in institutions. These results highlight the importance of structural factors for institutional trust, in line with the performance theory of trust, and are consistent with prior research. Research has shown that lower parental socio-economic status at birth was prospectively associated with less trust in political institutions at ages 30-33 (Schoon & Cheng, 2011), and other studies have documented mistrust in institutions among racialized groups as a result of discrimination and structural inequality (Wilkes & Wu, 2018). School difficulties may also lead to structural disadvantage: schools may not adequately meet the needs of adolescents with learning difficulties, and the resulting lower performance in school can have a lasting impact on educational attainment and socio-economic status.

However, the causal effects of socio-economic and racialized groups on institutional trust were not supported by the tPC algorithm. It is likely that the mechanisms linking these factors to institutional trust are heterogeneous, complicating their identification at the population level as in the present study. For example, in some individuals, low household income at birth may impact future trust in institutions through lower access to educational and occupational resources (Schoon & Cheng, 2011), whereas in other individuals, the key mechanism may be the adverse experience of living in an unsafe neighborhood (Intravia et al., 2016). Mutual adjustment for household income and racialized minority status in the tPC algorithm might have occulted their shared influences on structural inequality and institutional trust. In addition, different racialized groups will have varied experiences of discrimination and social inclusion, which we could not parse out here due to the small number of individuals within groups other than White. Given this unexplored heterogeneity, it may be necessary to employ more granular measurements, including using qualitative and mixed methods, to better understand how specific groups and their intersections develop low trust in institutions.

The putative effect of interpersonal factors on institutional trust was supported, to some extent, by the associations of bullying exposure with less institutional trust, and of social capital with greater institutional trust. Other risk factors identified in the regression models (but not in the tPC algorithm) were male sex and maternal mental health problems. Sex or gender differences in institutional trust have not been consistent in the literature (Xiao & McCright, 2015) but may echo, here, the polarization of socially disenfranchised young men (e.g., the incel phenomenon) (Broyd et al., 2023). Maternal mental health problems may be related to institutional trust through confounding influences (e.g., lower income) or via effects on youth’s mental health and the parent-child relationship. The absence of association with paternal mental health could be due to the greater proportion of missing data or lower involvement of fathers in some households.

Associations were not significant for neighborhood trust, family cohesion, gender diverse status, and social media use. Both family cohesion and neighborhood trust at age 15 were reported by mothers rather than by participants. It is possible that these external accounts did not reliably capture participants’ experiences of social connectedness during adolescence. Another consideration is that during adolescence, factors that influence relationships with peers in the school environment (such as bullying and mental health problems) might be more influential on youth’s sense of belonging and trust, compared to experiences in other spheres of their environment (such as family and neighborhood) – pointing to the role of school as the primary institution during adolescence. The lack of association of gender diverse status with institutional trust may be the product of insufficient statistical power due to small group size, as well as because of heterogenous experiences of interactions with institutions, as we suggested above in relation to racialized minority status. The lack of association for social media use may similarly be due to heterogeneous causal pathways that compete with each other at the group level: social media might amplify or alleviate mistrust depending on how the technology is used (Paquin, Ackerman, et al., 2024), and its use may be influenced by prior mental health problems and structural factors (Houghton et al., 2018; Nagata et al., 2022; Paquin, Philippe, et al., 2024).

### Mental health and institutional trust

To our knowledge, this is the first study examining prospective associations of mental health problems with institutional trust, confirming prior cross-sectional findings in adults (Buck-McFadyen et al., 2019; Economou et al., 2014; Hudson, 2006; Lindstrom & Mohseni, 2009). In examining associations between mental health and institutional trust prospectively, we found that internalizing and externalizing problems at age 15 years were associated with less institutional trust at age 23, independent of early-life factors. This result suggests that adolescent mental health problems are a potential risk factor for lower levels of trust in institutions.

Mental health problems during adolescence may contribute to lower levels of institutional trust through a combination of social and psychological mechanisms. In the results of the tPC algorithm, externalizing (but not internalizing) problems had indirect associations with institutional trust through their putative effects on psychotic experiences. Internalizing and externalizing problems are known risk factors for psychotic experiences (e.g., Healy et al., 2020), and general mistrust has been proposed to be an intermediary phenotype en route to the emergence of psychotic experiences (Catone et al., 2024). Mental health problems may contribute to institutional mistrust through stigma and social exclusion in the school environment: adolescents experiencing mental health problems, and particularly externalizing difficulties, are at increased risk of stigma and rejection by their peers (Corrigan et al., 2005; Lau et al., 2016; Martin et al., 2007). Social exclusion may in turn foster paranoid thinking (Bell et al., 2023), as captured here by the measure of psychotic experiences at age 23, eventually leading to poorer institutional trust. The role of social exclusion in these associations was partially supported by the (undirected) edge between externalizing problems and exposure to bullying, and the edge between social capital and institutional trust, whose unstable orientation could reflect a bidirectional relationship.

An alternative interpretation is that mental health problems in adolescence are not causally related to institutional mistrust and are instead related as markers of risk. This non-causal explanation cannot be ruled out: although the tPC aims to identify causal relationships, it makes strong assumptions that can hardly be met in social sciences, notably that all confounding factors were included in the set of considered variables. For instance, one of the potential confounders not included here is genetic predisposition (Sturgis et al., 2010), which could hypothetically be a shared risk factor for institutional mistrust and mental health problems.

Higher levels of mental health problems at age 23 were also associated cross-sectionally with less trust in institutions at age 23, and the association between perceived stress and institutional trust was oriented in the reverse direction by the tPC algorithm, meaning that the lack of institutional trust putatively contributed to more stress in adulthood. This suggested effect of mistrust on stress could reflect the psychological strain of unreliable, unfair, or hostile treatment from institutions (C. Morgan et al., 2019).

### Implications of the findings

Although further research on developmental pathways towards institutional trust is needed, the results point to the interplay of social and mental health factors across developmental stages, underlining the importance of a developmental and systemic perspective on the study of institutional trust. The improvement of social conditions during early life has the potential to foster better trust in institutions over the life course. Because adolescents with mental health problems tend to develop less trust in institutions, youth mental health services may also be an appropriate space to intervene on trust, especially given the potential reciprocal effects of mistrust on mental health problems. However, considering the interplay of psychological and social factors shaping trust, interventions that focus on psychological factors during adolescence or adulthood without addressing the social dimension would likely be insufficient. Social interventions may help restore trust in institutions if they reinforce the young person’s sense of belonging within society and demonstrate that institutions are successfully enabling them to thrive within it. For instance, approaches that involve shared activities with others, such as sports- or arts-based interventions, can help individuals expand their social capital within and beyond their local communities (H. Morgan & Parker, 2017), with hypothetical benefits for replenishing institutional trust. In sum, a comprehensive agenda for improving institutional trust should propose interventions that span life stages and the psychological and social domains.

### Strengths and limitations

This study benefited from prospective data covering socio-demographic, household, parental, interpersonal, and mental health characteristics over 23 years. By combining traditional regression analyses with causal discovery approaches, we identified risk factors for institutional mistrust and possible mechanistic pathways. An important limitation of the study, however, is the potential for sampling bias. Families were recruited using recruitment material in English or French, limiting access for families who did not understand those languages, and those residing in Indigenous communities were not included. Families with less trust in institutions may also have been more likely to decline study participation or to withdraw from the study before age 23. As a result, the results may not generalize well to the populations that are the least trusting of institutions. Trust was likely affected by how participants perceived institutional responses to the COVID-19 pandemic: institutional trust was measured between February and June 2021, when the state of emergency was ongoing. Lastly, relevant measures, such as the contrast between in-group and out-group relationships (sometimes referred to as “bonding” and “bridging” social capital; Claridge, 2018) were not available.

Of note, the tPC algorithm cannot identify cyclic associations between two variables, such as a bidirectional relationship where social capital and institutional trust at age 23 mutually influence each other. In practice, it seems likely that many variables had bidirectional relationships; these had to be collapsed by the algorithm into a single direction or left undirected. Identification of bidirectional relationships ideally requires repeated measures of both variables, and to our knowledge very limited data of the sort have been published in relation to institutional trust (e.g., Brehm & Rahn, 1997). The present findings should therefore be interpreted with caution given the possibility of unidentified bidirectional relationships, which may partly explain why bootstrap analyses often failed to show consistent directions of association.

## Conclusion

This study found longitudinal associations of social and mental health factors, from early life to adulthood, with institutional trust. The results suggest that the chains of causation leading to institutional trust may contain an interplay of factors related to structural inequality, interpersonal factors, and mental health, but these pathways likely differ depending on the unique context of each individual. Future research should consider how structural and mental health interventions can support greater social inclusion across populations to improve institutional trust and wellbeing.

## DATA ACCESS

Participants of the Québec Longitudinal Study of Child Development only consented to share their data with the study’s financial partners and affiliated researchers and their collaborators. To access data from the cohort, researchers should send a request to the Institut de la statistique du Québec (https://statistique.quebec.ca/en/institut/services-for-researchers).

