## Supplementary Material for "Social and Mental Health Pathways to Institutional Trust: A Cohort Study"

### Note S1. Factor analyses of institutional trust

We began factor analyses by randomly splitting the total analytical sample into two independent subsamples of equal sizes. We used the first subsample for exploratory factor analyses and the second subsample for confirmatory factor analyses. All models were estimated using lavaan version 0.6-17 with weighted least squares mean and variance adjusted methods.^1^ Exploratory factor analyses were done with oblimin oblique rotation. We selected the number of factors according to the scree plot and eigenvalues. Items with no factor loading >0.4 were removed.^2^ For confirmatory factor analyses, we used robust, mean and variance adjusted fit statistics. We defined comparative fit index (CFI)>0.90 as acceptable and >0.95 as good fit; and root mean square error approximation (RMSEA) and standardized root mean squared residual (SRMR) <0.08 as acceptable and <0.05 as good fit.^3,4^ If fit was not satisfactory, we revised the model based on modification indices and theoretical considerations.

The Kaiser-Meyer-Olkin was 0.89 and the Bartlett’s test for sphericity was significant (p<.001), supporting the suitability of the data for factor analysis. The eigenvalues and scree plot indicated a 3-factor solution, which we explored in the first subsample (n=673; Figure A1). One item (local merchants) had no primary factor loading above 0.40 and was thus excluded in subsequent analyses and from the total trust score.

**Figure A1.** Exploratory factor analysis (n=673) with 3 factors

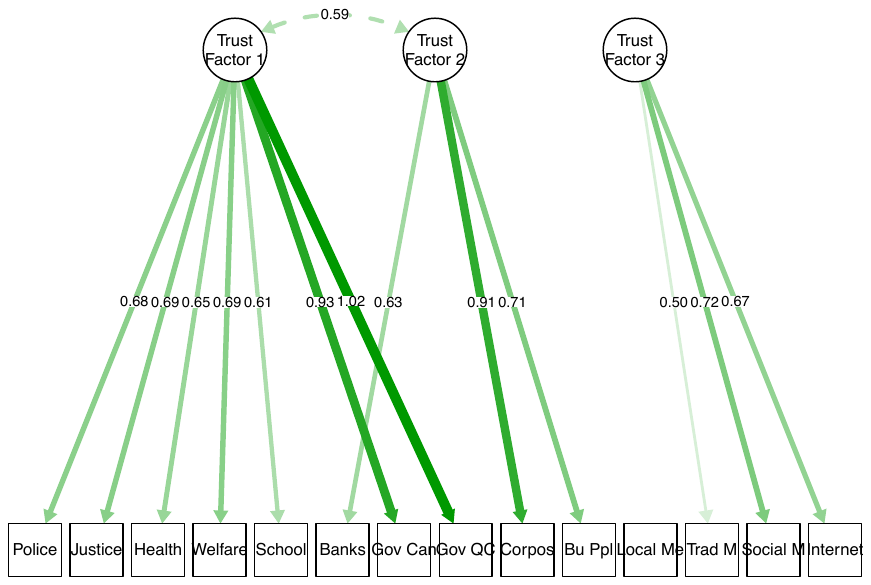

From left to right, items are the police, justice system and courts, health care system, welfare system, school system, banks, federal parliament (of Canada), provincial parliament (of Québec), major corporations, businesspeople, local merchants, traditional media, social media, and internet. Data compiled from the Québec Longitudinal Study of Child Development (1998–2021), Gouvernement du Québec, Institut de la statistique du Québec.

In the confirmatory analysis, the fit of this 3-factor model was not satisfactory: χ^2^(62)=763.70, *p*<.001; CFI=0.959; RMSEA=0.130 (90% CI: 0.122, 0.138); SRMR=0.067. Based on modification indices, we added residual correlations between police and justice, federal and provincial parliaments, banks and businesspeople, health care and school, welfare and school, and welfare and health care. The resulting 3-factor solution achieved acceptable to good fit: χ^2^(56)=270.87, *p*<.001; CFI=0.987; RMSEA=0.076 (90% CI: 0.067, 0.085); SRMR=0.047. We labeled the factors as trust in social, economic, and mediatic institutions (Figure 1 in the main text). Internal reliability was adequate: respectively ω=0.827, ω=0.860, and ω=0.714.

### Note S2. Selection of social and mental health variables

The Québec Longitudinal Study of Child Development has been collecting data every 1 to 2 years since 1997, with 26 time points to date and several thousands of variables. This presented us with the dilemma of selecting which measures and time points to include in the present study. Below, we outline our decision process that led to the selection of variables presented in the article.

***General considerations***

We were broadly interested in the role of social and mental health factors, from birth to early adulthood, on institutional trust. We heuristically categorized these factors according to three categories of determinants, based on theorized causal mechanisms: structural inequality, interpersonal factors, and mental health (see the article introduction). In relation to these three categories, we additionally aimed to account for key antecedents or confounding factors: e.g., parental history of mental health problems as a shared risk factor for mental health and social difficulties of participants.

In selecting social and mental health variables, we aimed to balance comprehensiveness and parsimony, focusing on the factors that appeared most salient based on theory. We therefore constrained the selection of measures to 3 time points between birth and adulthood in order to simplify the analyses, their interpretation, and their reporting.

***Selection of time points***

*Early life.* We measured socio-demographic factors and parental history of mental health problems in early life, given that their putative effects on structural inequality and institutional trust could potentially operate over the life course.^5^

*Adolescence*. We initially considered having the second time point in late childhood or pre-adolescence (e.g., age 12) as a midpoint between early life and adulthood. However, we ultimately opted for a time point in adolescence (at age 15) since mental health problems more commonly emerge during that period or later.^6^

*Adulthood.* Institutional trust was measured at age 23. We opted to include adulthood predictors at the same age (i.e., cross-sectionally) given the availability of key variables at this age, notably social capital and psychotic experiences. An additional advantage of having predictors that were cross-sectional to institutional trust (rather than anterior) was the possibility of identifying reverse associations (e.g., from trust to mental health) in the temporal Peter-Clark algorithm.

**Measures related to structural inequality**

Measures related to structural inequality were racialized minority status, gender diverse status, low household income, and school difficulties. We included racialized minority status and gender diverse status based on their risk of exposure to discrimination and lower trust in institutions.^7,8^ We measured gender diverse status at age 23 given that the realization of gender diverse identity often occurs during adolescence or later.^9^ Lower socio-economic status (indexed here with household income) can engender unfair treatment from institutions and is commonly associated with lower trust in institutions.^10^ We included school difficulties because, even though they are not typically described as a source of structural inequality, they can have lasting effects on wealth and social disparities that are consistent with the definition of structural inequality employed here.^11^

**Measures related to interpersonal relationships**

Measures related to interpersonal relationships included neighborhood trust, family cohesion, exposure to bullying, school difficulties, social capital, and social media use. The rationale for these inclusions is that neighborhood trust^12^ and family cohesion^13^ contribute to social connectedness, whereas school difficulties and bullying may hamper the sense of belonging in the school environment.^14^ Interpersonal relationships and social capital are closely related:^15^ social capital is notably defined by the availability of supportive and reciprocal relationships. We included social media use as a factor variably related to social connections, in the sense that social media can foster social connections or undermine them, depending on the types and contexts of use;^16^ and social media may further influence trust in institutions by disseminating information that makes institutions appear less trustworthy.^17^

**Measures related to mental health**

We did not have strong *a priori* to anticipate that any particular category of mental health problems would be more strongly related to institutional trust than others. One exception is the spectrum of psychotic experiences, which includes paranoia and has been linked to interpersonal mistrust;^18^ although the link of psychotic experiences with institutional trust has not been previously examined, we expected that its putative impact on interpersonal trust would also lead to lower trust in institutions. For other measures related to mental health, we selected transdiagnostic markers of mental health problems, spanning all levels of severity. In adolescence, we included internalizing and externalizing problems. In adulthood, we included perceived stress (a sensitive marker at low levels of psychopathology), psychotic experiences, and two markers of more severe mental health problems: help seeking and suicidal ideations.

### Table S1. Description of mental health and social measures

| **Measure** | **Informant (age)** | **Content** | **Coding** |
| --- | --- | --- | --- |
| ***Early life*** | | | |
| Sex | Medical records (birth) | Male or female | 0 = Male  1 = Female |
| Racialized minority status | Parents (5 months) | 12 categories, e.g., “White”, “Black” | 0 = “White”  1 = any of the other categories |
| Household income insufficiency | Parents (5 months) | Defined as exceeding by 20% or more the average proportion of annual income spent on basic needs among households of similar size and regional population density ^19^. | 0 = Sufficient  1 = Insufficient |
| Parental history of depression | Parents (29 months) | 16 items from the Diagnostic Interview Schedule, e.g., periods of feeling sad most of the time for 2 weeks or more ^20^ | 0 = No history of depression  1 = History of depression |
| Parental antisocial behaviors in adolescence | Parents (5 months) | 5 items from the Diagnostic Interview Schedule, e.g., stealing, skipping school ^21^ | Sum score, range: 0–5 |
| ***Adolescence*** | | | |
| Neighborhood trust | Mothers (15 years) | 5 items, ad hoc, e.g., people help each other, adults can be trusted ^22^ | Sum score, range: 1-4 |
| Family cohesion | Mothers (15 years) | 7 items from the McMaster Family Assessment Device, e.g., “We confide in each other”, “We feel accepted for what we are” ^23,24^ | Sum score, range: 7-28 |
| Bullying exposure | Self (15 years) | 6 items adapted from the Self-Report Victimization Scale, e.g., being called names, getting pushed, hit or kicked ^25^, | Sum score: range: 6-18 |
| School difficulties | Self (15 years) | Dropout Prediction Index, 8 items, e.g., average grades, motivation for school ^26^ | Formula (see reference), range: 0-1 |
| Internalizing problems | Self (15 years) | 25 items from the Mental Health and Social Inadaptation Assessment for Adolescents ^27^ | Sum score, rescaled to range: 0-10 |
| Externalizing problems | Self (15 years) | 57 items from the Mental Health and Social Inadaptation Assessment for Adolescents ^27^ | Sum score, rescaled to range: 0-10 |
| ***Adulthood*** | | | |
| Gender diverse status | Self (23 years) | Gender assessed with 1 item (options: masculine, feminine, or other) and coded in relation to sex assigned at birth. | 0 = cisgender  1 = transgender or other gender |
| Perceived stress | Self (23 years) | Perceived Stress Scale – Past-month version, 10 items, e.g., feeling nervous and stressed, feeling on top of things (reverse coded) ^28^ | Sum score, range: 0-40 |
| Psychotic experiences | Self (23 years) | Community Assessment of Psychic Experiences – Lifetime version, 15 items, e.g., “Have you ever felt as if the thoughts in your head are not your own?” ^29^ | Sum score, range: 15-60 |
| Suicidal ideas | Self (23 years) | 1 item: “In the past 12 months, did you ever seriously consider taking your own life or killing yourself?” | 0 = No  1 = Yes |
| Help seeking | Self (23 years) | 1 item: “In the past 12 months, have you sought professional help for psychological difficulties such as loneliness, sadness, anxiety, depression, violence or substance abuse?” | 0 = No  1 = Yes |
| Social capital | Self (23 years) | Social Provisions Questionnaire, 10 items, e.g., “There are people I can depend on”, “I have close relationships” ^30^ | Sum score, range: 0-40 |
| Social media use | Self (23 years) | 1 item: “During the past 3 months, how much time on average per day did you spend using social media such as Facebook, Instagram, Snapchat, Twitter or TikTok?” (options ranging from 0 = “did not use” to 6 = “6 hours or more”) ^31^ | Transformed into numeric values based on the midpoint of response categories: 0, 0.5, 2, 4.5, and 6 |

### Table S2. Data availability in the analytic sample (n=1347)

| **Variable** | **Availability, N** | **Availability, %** |
| --- | --- | --- |
| Sex assigned at birth | 1347 | 100 |
| Racialized group | 1344 | 99.8 |
| Household income | 1332 | 98.9 |
| Maternal history of depression | 1329 | 98.7 |
| Paternal history of depression | 1108 | 82.3 |
| Maternal antisocial behaviors in adolescence | 1308 | 97.1 |
| Paternal antisocial behaviors in adolescence | 1173 | 87.1 |
| Neighborhood trust | 1129 | 83.8 |
| Family cohesion | 968 | 71.9 |
| Bullying exposure | 1197 | 88.9 |
| School difficulties | 1188 | 88.2 |
| Internalizing behaviors | 1199 | 89.0 |
| Externalizing behaviors | 1199 | 89.0 |
| Gender minority status | 1347 | 100.0 |
| Perceived stress | 1345 | 99.9 |
| Psychotic experiences | 1316 | 97.7 |
| Suicidal ideas | 1321 | 98.1 |
| Help seeking | 1347 | 100.0 |
| Social capital | 1347 | 100.0 |
| Social media use | 1347 | 100.0 |
| Institutional trust | 1347 | 100.0 |

### Table S3. Early-life characteristics of participants included in and excluded from the analyses

|  | **Participant, No. (%)** | | **Effect size** |
| --- | --- | --- | --- |
|  | **Included**  **N=1347** | **Excluded**  **N=773** |  |
| Sex assigned at birth |  |  | 0.216 |
| Male | 576 (42.8%) | 504 (65.2%) |  |
| Female | 771 (57.2%) | 269 (34.8%) |  |
| Racialized minority group |  |  | 0.094 |
| Yes | 110 (8.18%) | 109 (14.1%) |  |
| No | 1234 (91.8%) | 664 (85.9%) |  |
| Low household income at 5 months^a^ |  |  | 0.135 |
| Yes | 1063 (79.8%) | 508 (67.7%) |  |
| No | 269 (20.2%) | 242 (32.3%) |  |
| Single parent household at 5 months |  |  | 0.071 |
| Yes | 89 (6.63%) | 82 (10.7%) |  |
| No | 1254 (93.4%) | 687 (89.3%) |  |
| Maternal age in years at child’s birth, Mean (SD) | 29.2 (5.10) | 28.4 (5.40) | 0.071 |
| Maternal educational attainment at 5 months |  |  | 0.110 |
| No high school diploma | 309 (23.0%) | 255 (33.1%) |  |
| High school diploma or higher | 1037 (77.0%) | 516 (66.9%) |  |
| Maternal depressive symptoms at 5 months, Mean (SD)^b^ | 1.33 (1.28) | 1.54 (1.43) | 0.070 |
| Internalizing behaviors at 29 months, Mean (SD)^c^ | 1.15 (0.20) | 1.15 (0.21) | 0.004 |
| Externalizing behaviors at 29 months, Mean (SD)^c^ | 1.50 (0.29) | 1.51 (0.31) | 0.011 |

Effect sizes for comparing included and excluded participants are Cramer’s V and absolute Spearman correlations (range: 0.000–1.000). Reasons for exclusion were no assessment at age 23 (n=752) or missing data on institutional trust (n=21). Data compiled from the Québec Longitudinal Study of Child Development (1998–2021), Gouvernement du Québec, Institut de la statistique du Québec.

^a^ Low income defined as spending more than 20% of the annual income for basic needs, in addition to the average proportion spent by households of similar size and regional population density.

^b^ 12-item version of the Center for Epidemiologic Studies-Depression,^32^ rescaled to range 0–10.

^c^ From the Behavior Questionnaire;^33^ mean scores (range 1-3) of 6 items for internalizing and 10 items for externalizing behaviors; missing values replaced with 17 months.

### Table S4. Edges among adulthood variables over 100 bootstraps of the temporal Peter-Clark algorithm (sparsity level=0.00001)

|  | **Gender minority** | **Perceived stress** | **Psychotic experiences** | **Suicidal ideas** | **Help seeking** | **Social capital** | **Social media use** | **Institutional trust** |
| --- | --- | --- | --- | --- | --- | --- | --- | --- |
| **Gender minority** | 0 (0) | 0 (0) | 0 (0) | 0 (0) | 0 (0) | 0 (0) | 0 (0) | 0 (0) |
| **Perceived stress** | 0 (0) | 0 (0) | 91 (0) | 40 (0) | 96 (0) | 61 (0) | 0 (0) | 44 (0) |
| **Psychotic**  **experiences** | 0 (0) | 8 (0) | 0 (0) | 1 (0) | 0 (0) | 13 (3) | 7 (0) | 28 (10) |
| **Suicidal ideas** | 0 (0) | 9 (0) | 0 (0) | 0 (0) | 9 (66) | 2 (0) | 0 (0) | 0 (0) |
| **Help seeking** | 0 (0) | 1 (0) | 0 (0) | 12 (66) | 0 (0) | 0 (0) | 0 (0) | 0 (0) |
| **Social capital** | 0 (0) | 0 (0) | 5 (3) | 1 (0) | 0 (0) | 0 (0) | 0 (0) | 10 (43) |
| **Social media use** | 0 (0) | 0 (0) | 0 (0) | 0 (0) | 0 (0) | 0 (0) | 0 (0) | 0 (0) |
| **Institutional trust** | 0 (0) | 0 (0) | 34 (10) | 0 (0) | 0 (0) | 24 (43) | 0 (0) | 0 (0) |

The numbers indicate how many times edges were identified over 100 bootstraps of the temporal Peter-Clark algorithm. The first number in each cell indicates bootstraps where the variable at the top (columns) had a putative causal effect on the variable on the left (rows). The second number in parentheses indicates bootstraps where the edge was undirected, that is, where the direction of causality was undetermined. Data compiled from the Québec Longitudinal Study of Child Development (1998–2021), Gouvernement du Québec, Institut de la statistique du Québec.

### Figure S1. Associations of early-life factors with institutional trust (adjusted for sex)

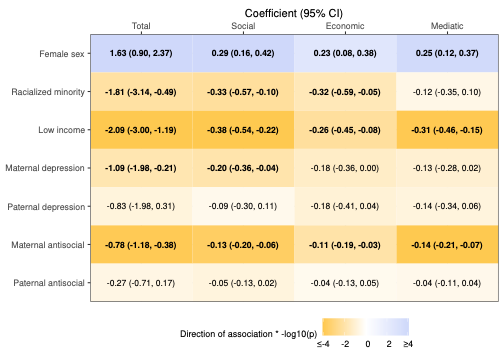
 Linear regression models pooled over 40 multiply imputed datasets (n=1347). Cells indicate unstandardized regression coefficients (95% confidence intervals). Each association is modeled separately. Bold indicates p<.05 after adjustment for false discovery rate across columns. Darker shades indicate smaller p-values (orange: negative association; blue: positive association). Data compiled from the Québec Longitudinal Study of Child Development (1998–2021), Gouvernement du Québec, Institut de la statistique du Québec.

### Figure S2. Associations of adolescence factors with institutional trust (adjusted for sex)

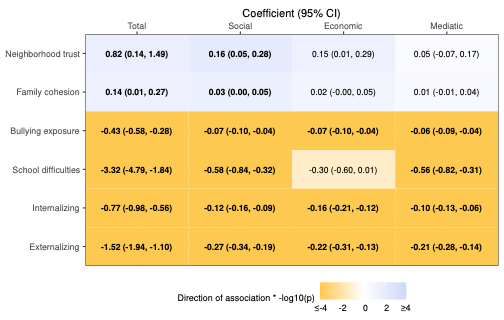
 Linear regression models pooled over 40 multiply imputed datasets (n=1347). Cells indicate unstandardized regression coefficients (95% confidence intervals). Each association is modeled separately. Bold indicates p<.05 after adjustment for false discovery rate across columns. Darker shades indicate smaller p-values (orange: negative association; blue: positive association). Data compiled from the Québec Longitudinal Study of Child Development (1998–2021), Gouvernement du Québec, Institut de la statistique du Québec.

### Figure S3. Associations of adolescence factors with institutional trust (adjusted for sex and early-life factors)

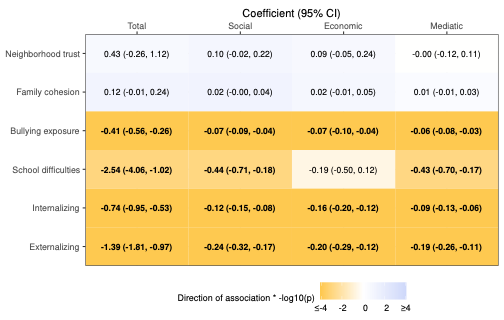
 Linear regression models pooled over 40 multiply imputed datasets (n=1347). Cells indicate unstandardized regression coefficients (95% confidence intervals). Each association is modeled separately. Bold indicates p<.05 after adjustment for false discovery rate across columns. Darker shades indicate smaller p-values (orange: negative association; blue: positive association). Data compiled from the Québec Longitudinal Study of Child Development (1998–2021), Gouvernement du Québec, Institut de la statistique du Québec.

### Figure S4. Associations of adulthood factors with institutional trust (adjusted for sex)

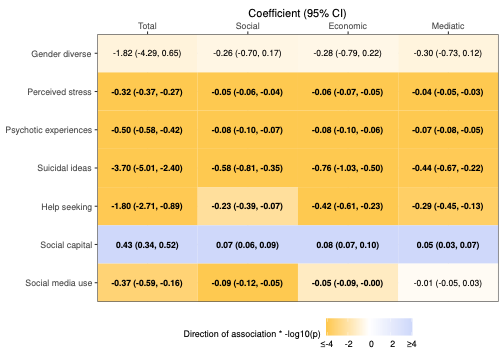
 Linear regression models pooled over 40 multiply imputed datasets (n=1347). Cells indicate unstandardized regression coefficients (95% confidence intervals). Each association is modeled separately. Bold indicates p<.05 after adjustment for false discovery rate across columns. Darker shades indicate smaller p-values (orange: negative association; blue: positive association). Data compiled from the Québec Longitudinal Study of Child Development (1998–2021), Gouvernement du Québec, Institut de la statistique du Québec.

### Figure S5. Associations of adulthood factors with institutional trust (adjusted for sex and early-life factors)

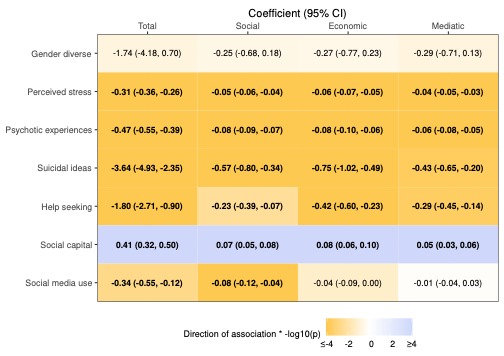
 Linear regression models pooled over 40 multiply imputed datasets (n=1347). Cells indicate unstandardized regression coefficients (95% confidence intervals). Each association is modeled separately. Bold indicates p<.05 after adjustment for false discovery rate across columns. Darker shades indicate smaller p-values (orange: negative association; blue: positive association). Data compiled from the Québec Longitudinal Study of Child Development (1998–2021), Gouvernement du Québec, Institut de la statistique du Québec.

### Figure S6. Associations of adulthood factors with institutional trust (adjusted for sex and adolescence factors)

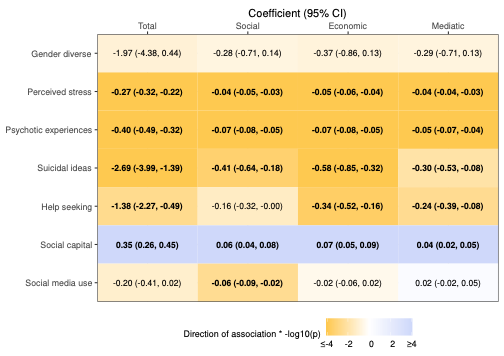
 Linear regression models pooled over 40 multiply imputed datasets (n=1347). Cells indicate unstandardized regression coefficients (95% confidence intervals). Each association is modeled separately. Bold indicates p<.05 after adjustment for false discovery rate across columns. Darker shades indicate smaller p-values (orange: negative association; blue: positive association). Data compiled from the Québec Longitudinal Study of Child Development (1998–2021), Gouvernement du Québec, Institut de la statistique du Québec.

### Figure S7. Pathways between mental health and social factors and institutional trust (sparsity=0.01)

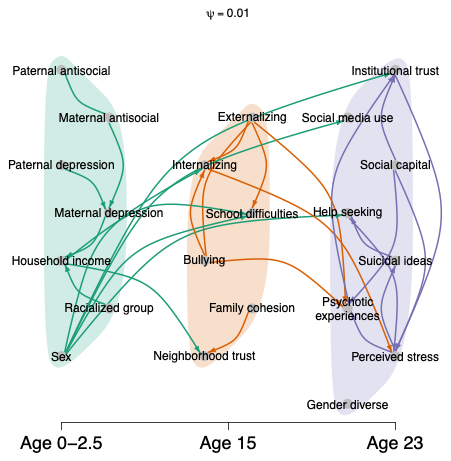

Temporal Peter-Clark algorithm with test-wide delection (n=1347). Edges (lines) indicate putative causal relationships between variables. Some edges have no arrowheads, indicating that the direction of causality could not be determined by the algorithm. Data compiled from the Québec Longitudinal Study of Child Development (1998–2021), Gouvernement du Québec, Institut de la statistique du Québec.

### Figure S8. Pathways between mental health and social factors and institutional trust (sparsity=0.001)

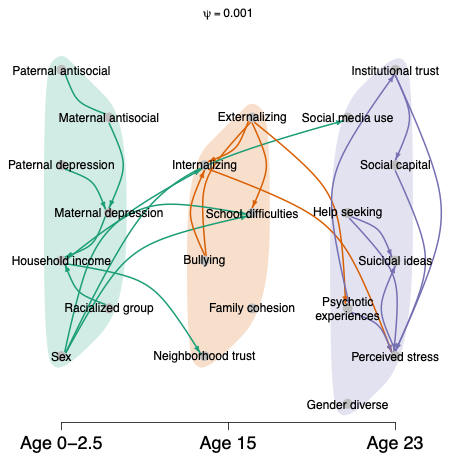

Temporal Peter-Clark algorithm with test-wide delection (n=1347). Edges (lines) indicate putative causal relationships between variables. Some edges have no arrowheads, indicating that the direction of causality could not be determined by the algorithm. Data compiled from the Québec Longitudinal Study of Child Development (1998–2021), Gouvernement du Québec, Institut de la statistique du Québec.

### Figure S9. Pathways between mental health and social factors and institutional trust (sparsity=0.0001)

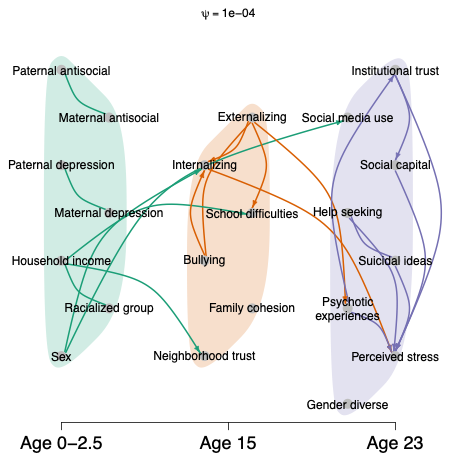

Temporal Peter-Clark algorithm with test-wide delection (n=1347). Edges (lines) indicate putative causal relationships between variables. Some edges have no arrowheads, indicating that the direction of causality could not be determined by the algorithm. Data compiled from the Québec Longitudinal Study of Child Development (1998–2021), Gouvernement du Québec, Institut de la statistique du Québec.
